# Impact of prior SARS-CoV-2 infection on post-vaccination SARS-CoV-2 spike IgG antibodies in a longitudinal cohort of healthcare workers

**DOI:** 10.1101/2021.09.16.21263576

**Authors:** Diana Zhong, Shaoming Xiao, Amanda K. Debes, Emily R. Egbert, Patrizio Caturegli, Elizabeth Colantuoni, Aaron M. Milstone

## Abstract

Waning serum antibodies against SARS-CoV-2 have sparked discussions about long-term immunity and need for vaccine boosters. We examined SARS-CoV-2 spike IgG antibodies in a longitudinal cohort, comparing antibody decay in individuals who received an mRNA SARS-CoV-2 vaccine, with and without prior SARS-CoV-2 infection. We completed a longitudinal cohort of healthcare workers (HWs) between June 2020 and September 2021. HWs were included if they had a serum sample collected after SARS-CoV-2 infection and/or a serum sample collected ≥ 14 days after second dose of an mRNA SARS-CoV-2 vaccine. Linear regression models adjusting for vaccine type, age, and sex were used to compare post-vaccination antibody levels between 1) HWs with and without prior SARS-CoV-2 infection and 2) HWs with prior SARS-CoV-2 infection ≤ 90 days and > 90 days prior to first vaccine. Serum was collected from 98 HWs after SARS-CoV-2 infection and before vaccine, and 1960 HWs ≥ 14 days following second vaccine dose. Serum spike antibody levels were higher after vaccination than after natural infection. Compared to SARS-CoV-2 naïve individuals, those with prior infection maintained higher post-vaccination mean spike IgG values at 1, 3, and 6 months, after adjusting for age, sex, and vaccine type. Individuals with PCR-confirmed infection > 90 days before vaccination had higher post-vaccination antibody levels than individuals infected ≤ 90 days before vaccination. Individuals with three exposures to spike protein maintain the highest antibody levels particularly when first and second exposures were greater than 90 days apart. A booster dose provides a third exposure and may similarly induce a more durable antibody response.

## Introduction

Waning serum antibodies against SARS-CoV-2 have sparked intense public health interest about long-term immunity. Lower antibody levels to SARS-CoV-2 spike protein are associated with breakthrough infections after vaccination, prompting consideration of booster doses.^1,2^ Prior infection may enhance protection from vaccination, stimulating inquiry about the impacts of hybrid immunity. Our objective was to examine SARS-CoV-2 spike IgG antibodies in a longitudinal cohort, comparing antibody decay in individuals who received an mRNA SARS-CoV-2 vaccine, with and without prior SARS-CoV-2 infection.

## Methods

Healthcare workers (HWs) were consented and enrolled starting June 3, 2020 and followed through September 3, 2021. HWs provided serum samples longitudinally, separated by at least 90 days. SARS-CoV-2 PCR results and vaccination dates were collected from electronic health records. HWs were included if they had a serum sample collected after SARS-CoV-2 infection (defined as positive SARS-CoV-2 PCR), and/or a serum sample collected ≥ 14 days after second dose of an mRNA SARS-CoV-2 vaccine. Serum was tested using an enzyme-linked immunosorbent assay (ELISA) [Euroimmun], as previously described.^3,4^ Linear regression models adjusting for vaccine type, age, and sex were used to compare post-vaccination antibody levels (log-transformed) between 1) HWs with and without prior SARS-CoV-2 infection and 2) HWs with prior SARS-CoV-2 infection ≤ 90 days and > 90 days prior to first vaccine. Ethical approval was obtained from the Johns Hopkins University Institutional Review Board. Analyses were performed in R, version 4.0.2.

## Results

Of 2009 included HWs, serum was collected from 98 HWs (143 samples) after SARS-CoV-2 infection and before vaccine, and 1960 HWs (2315 samples) ≥ 14 days following second vaccine dose (49 contributed to both groups). Of the 2009 HWs, 80% were female. 95% were Non-Hispanic/Latino and 80% were White. The median age was 40.4 years (IQR: 32.6, 52.1).

Serum spike antibody levels were higher after vaccination than after natural infection (Figure 1a). Compared to SARS-CoV-2 naïve individuals, those with prior infection maintained higher post-vaccination mean spike IgG values by 14, 19 and 56 percent at 1, 3, and 6 months, respectively, after adjusting for age, sex, and vaccine type (Table 1). Individuals with PCR-confirmed infection > 90 days before vaccination had higher post-vaccination antibody levels than individuals infected ≤ 90 days before vaccination by roughly 10 percent, at 1 and 3 months (Figure 1b).

**Figure 1.**
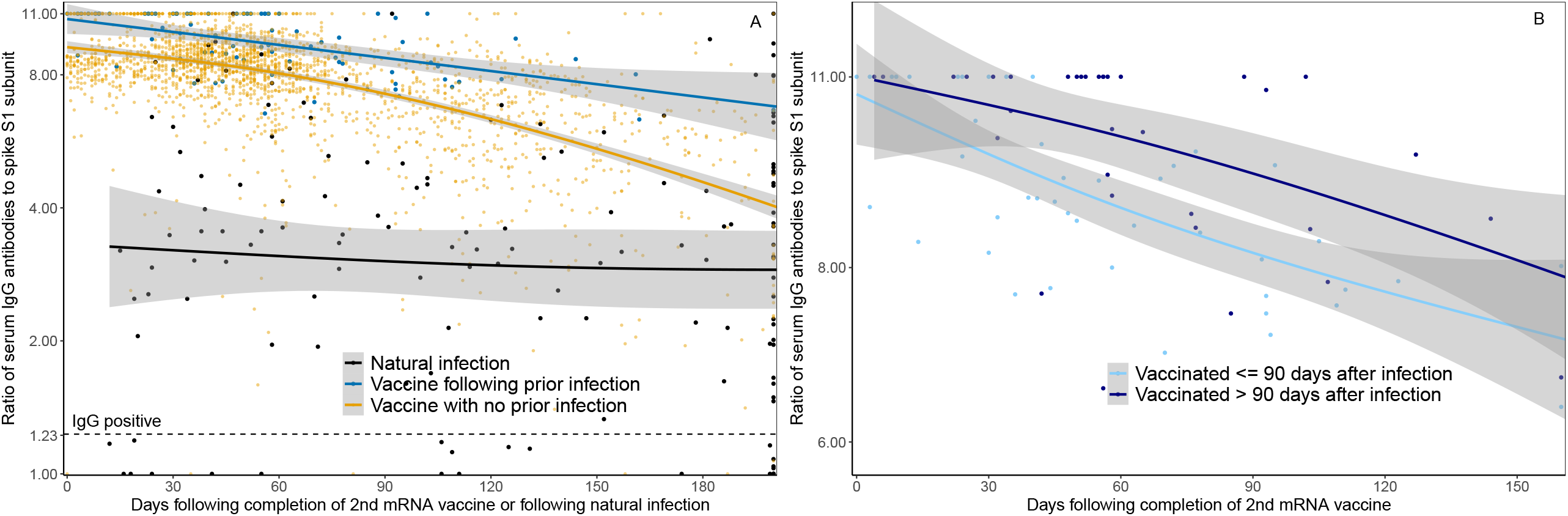
Waning IgG antibodies to SARS-CoV-2 following infection and vaccination in HWs: Panel A demonstrates serum spike S1 IgG antibody measurements in healthcare workers ≥ 14 days following dose two of SARS-CoV-2 mRNA vaccine with prior SARS-CoV-2 infection (blue line), ≥ 14 days following dose two of SARS-CoV-2 mRNA vaccine in those without prior SARS-CoV-2 infection (orange line), and in those following SARS-CoV-2 positive PCR test and before vaccination (black line). Prior SARS-CoV-2 infection was defined as positive SARS-CoV-2 PCR prior to first dose of mRNA vaccine. The x-axis represents days of follow-up from infection or vaccination, rather than calendar time, such that 49 HWs contributed to both natural infection and post-vaccination groups, respectively. The lines represent median IgG as a function of days following mRNA vaccination or natural infection, based on natural cubic splines (2 degrees of freedom) for each group. Shaded areas represent 95% Confidence Intervals. Panel B illustrates serum spike S1 IgG antibody decay after mRNA vaccination among participants with SARS-CoV-2 infection ≤ 90 days before vaccination (light blue line) and > 90 days before vaccination (dark blue line). The lines represent median IgG following mRNA vaccination over time using natural cubic splines (2 degrees of freedom). Shaded areas represent 95% Confidence Intervals.

**Table 1.** Characteristics of 1960 vaccinated healthcare workers and adjusted median spike IgG antibody measurements (95% CI^a^), and relative median antibody measurement (95% CI^a^) for those 1) with and without prior SARS-CoV-2 infection and 2) with prior SARS-CoV-2 infection ≤ 90 days and > 90 days prior to first vaccine dose

|  |  |  |  |  |  |  |  |
| --- | --- | --- | --- | --- | --- | --- | --- |
| Demographics | Infection prior to vaccine | No infection prior to vaccine |  |  | Infection prior to vaccine |  |  |
|  |  |  |  |  | Vaccinated > 90 days after infection | Vaccinated ≤ 90 days after infection |  |
| HWs, N | 73 | 1887 |  |  | 32 | 41 |  |
| Samples, N | 80 | 2235 |  |  | 34 | 46 |  |
| Age, median (IQR) | 37.1 (31.5, 48.2) | 40.3 (32.6, 52.1) |  |  | 41.0 (31.8, 50.5) | 35.9 (30.3, 47.6) |  |
| Male sex | 11(15) | 376 (20) |  |  | 5 (16) | 6 (15) |  |
| Vaccine type: Pfizer | 62 (85) | 1530 (81) |  |  | 28 (88) | 34 (83) |  |
| Time after vaccination | Adjusted Median IgG (95% CI) | Adjusted Median IgG (95% CI) | Relative Median (95% CI) |  | Adjusted Median IgG (95% CI) | Adjusted Median IgG (95% CI) | Relative Median (95% CI) |
| 1-month | 9.94 (9.58, 10.29) | 8.69 (8.56, 8.80) | 1.14(1.10, 1.19) |  | 10.52 (10.13, 11.00) | 9.66 (9.24, 10.02) | 1.09 (1.03, 1.16) |
| 3-months | 8.69 (8.27, 9.12) | 7.28 (7.15, 7.40) | 1.19 (1.13, 1.26) |  | 9.31 (8.47, 9.98) | 8.22 (7.81, 8.63) | 1.13 (1.02, 1.24) |
| 6-months<sup>b</sup> | 7.12 (6.29, 8.64) | 4.55 (4.16, 4.91) | 1.56 (1.35, 1.94) |  |  |  |  |

HW – healthcare workers, N – number, IQR – interquartile range
<sup>a</sup>Adjusted median IgG were estimated from linear regression models of log-transformed antibody level as a function of time (natural cubic spline with 2 degrees of freedom), group, interaction of time and group adjusting for vaccine type, age and sex. The 95% CIs for adjusted median IgG and relative median were constructed via the percentile bootstrap procedure using 1,000 bootstrap samples of HWs to account for clustering of serum samples within HWs.
<sup>b</sup>Adjusted median IgG and relative median at 6 months were not estimated for healthcare workers with prior SARS-CoV-2 infection ≤ 90 days and > 90 days prior to first vaccine dose separately due to few data points beyond 150 days.

## Discussion

Our study demonstrates that HWs who received two doses of mRNA vaccine maintain higher spike antibody levels than individuals with natural infection alone. HWs who had SARS-CoV-2 infection and then received two doses of mRNA vaccine (thus, three independent exposures to spike antigen) developed higher spike antibody levels than individuals with infection alone or those with vaccination alone, substantiating that vaccination after natural infection reduces risk of reinfection.^5^

Consistent with recent work comparing extended vaccine dosing intervals, a longer interval between infection and first vaccine dose enhances the antibody response.^6^ Individuals with three exposures to spike protein maintain the highest antibody levels particularly when first and second exposures were greater than 90 days apart. A booster dose provides a third exposure and may similarly induce a more durable antibody response. Limitations include defining SARS-CoV-2 infection as positive PCR, potentially misclassifying HWs with unconfirmed prior infection.

Further study is warranted to determine whether this increased post-vaccination antibody durability in previously infected individuals is attributable to number of exposures, interval between exposures, or a complex interplay between natural and vaccine-derived immunity. Given the prominent role of serum spike antibodies and virus neutralization ability as correlates of protection, and waning antibody levels in HWs, urgent studies are needed to determine how serological testing can inform optimal vaccine timing and the need for booster doses.

## Data Availability

Data are being analyzed as part of an ongoing cohort study. Data will be made available by request at the completion of the cohort study.

## Acknowledgements

The authors would like to thank members of the Johns Hopkins Hospital Clinical Immunology Laboratory, Danielle Koontz and Ani Voskertchian from the Johns Hopkins Division of Pediatric Infectious Diseases, Shaun Truelove from the Johns Hopkins Bloomberg School of Public Health, and Avinash Gadala from the Johns Hopkins Health System. Lastly, we thank Dr. Kirsten Vannice from the Bill and Melinda Gates Foundation for providing contribution to the analysis. Research reported in this publication was supported in part by the National Institute of Allergy and Infectious Diseases of the National Institutes of Health (NIH) under award number K24AI141580 (A.M.) and the generosity of the collective community of donors to the Johns Hopkins University School of Medicine and the Johns Hopkins Health System for Covid-19 research.

